# No woman should be left behind: A decomposition analysis of socioeconomic inequalities in unsafe abortion among women presenting for abortion care services in Lusaka and Copperbelt provinces of Zambia

**DOI:** 10.1101/2022.03.29.22273119

**Authors:** Patrick Kaonga, Moses Mukosha, Choolwe Jacobs, Margarate Nzala Munakampe, Victor Sichone, Christabel Chigwe Phiri, Musonda Makasa, Bellington Vwalika, Mwansa Ketty Lubeya

**Affiliations:** Department of Epidemiology and Biostatistics, School of Public Health, University of Zambia, Lusaka, Zambia; Department of International Health, Bloomberg School of Public Health, Johns Hopkins University, Baltimore 21218, Maryland, USA; Department of Pharmacy, School of Health Sciences, University of Zambia, Lusaka, Zambia; HIV and Women’s Health Research Group, University Teaching Hospital, Lusaka, Zambia; Department of Policy and Management, School of Public Health, University of Zambia, Lusaka, Zambia; Department of Obstetrics and Gynecology, Kitwe Teaching Hospital, Kitwe, Zambia; Zambia Association of Gynaecology and Obstetrics, Lusaka, Zambia; Young Emerging Scientists Zambia, Lusaka, Zambia; Department of Obstetrics and Gynecology, Levy Mwanawasa University Teaching Hospital, Lusaka, Zambia; Women and Newborn Hospital, University Teaching Hospitals, Lusaka, Zambia; Department of Obstetrics and Gynecology, School of Medicine, University of Zambia, Lusaka, Zambia

**Keywords:** Unsafe abortion, Women, Inequality, Concentration index, Decomposition, Zambia

## Abstract

This study measured socioeconomic-related unsafe abortion inequality among women presenting for abortion care services in Lusaka and the Copperbelt provinces of Zambia and decompose its causes. We conducted a cross-sectional study between August and September 2021. Unsafe abortion inequalities were assessed using corrected concentration index and Erreygers-type decomposition analysis was conducted to assess causes of unsafe abortion inequalities. Out of 362 women, the magnitude of unsafe abortion was 77(21.3%, [95% CI: 17.8, 24.9]). The corrected concentration index was -0.231 (95% CI: -0.309, -0.154), implying pro-poor inequality in unsafe abortion among women. Decomposition analysis showed that the major contributors of the unsafe abortion inequality were socioeconomic status (66.6%), marital status (6.3%), education (10.2%) and employment (3.7%). Also, history of unwanted pregnancy (5.1%), awareness of whether abortion is legal in Zambia (8.9%) and awareness that hospitals offered free abortion services (11.3%). The findings suggest that the unsafe abortion is a problem in Zambia and substantial inequality mainly due to socioeconomic factors. Stakeholders and policymakers should consider socioeconomic strategies to reduce unsafe abortion inequality promoting advocacy to increased access to legal safe abortion and use of modern contraceptives so that no woman is left behind in the prevention of unsafe abortion.

## Introduction

Unsafe abortion is a public health problem with a global average mortality rate estimated at 367 deaths per 100 000 which is unequally distributed, which two-fold higher in Africa. Developing countries bears 97% of the global unsafe abortion and in Africa 75% of all abortion are unsafe where as high as 50% of unsafe abortion may end up in complication with various morbidities [1-3] with about seven million women hospitalized yet all these consequences are preventable. The World Health Organization (WHO) defines an unsafe abortion as “the termination of pregnancy that is carried out by either an unskilled individual or in an environment that does not meet the minimal medical standards or both” [3].

There is mounting evidence that socioeconomic inequalities unequally distributed among individuals have a greater negative effect on those with lower socioeconomic status than those with higher socioeconomic status. There are disproportionate distribution in peripheral inputs such as residential area, wealth, education, employment and income contribute to the inequalities in health [4] and inputs are part of the common socioeconomic factors. One such important health concern has been unsafe abortion, which is a forgotten emergency and preventable problem. Unsafe abortion reflects one of the greatest health divides between rich and poor, and it is disproportionately distributed around the world and within countries suggest evidence that it is mainly pro-poor which is against the agenda for combating health inequalities [5, 6]. These variations may also reflect disparities in the pregnancy-related factors as provision of Comprehensive Abortion Care (CAC) services, accessibility and availability of CAC services [7]. Although increased access to legal abortion is associated with improved sexual and reproductive health outcomes [7], it is far from being a reality especially in sub-Saharan Africa where cultural, stigma and religious belief hinder many women from accessing CAC. This has contributed to women seeking abortion services from unqualified persons usually “wise women” or from environment that does not meet the medical standards [3].

It has been argued that health outcomes are more affected by differences in socioeconomic status than medical care. The relationship and inequalities between unsafe abortion and socioeconomic status is diverse among and within countries [8, 9] and lower socioeconomic status contribute to unsafe abortion. Thus, the variations in the magnitude and determinants in unsafe abortion across regions and within countries could be due to socioeconomic inequalities. Reports suggest that different socioeconomic levels among women contribute to the prevalence and distribution of unsafe abortion. Some studies have demonstrated importance of socioeconomic determinants such as lower socioeconomic status/income and other factors as major predictors of unsafe abortion [10, 11] which contributes to inequalities in unsafe abortion. In this regard, a study from Ghana showed that about 60% of women under 25 years were more likely to have unsafe abortion [12] and in other parts of Africa [13]. Similarly, other studies have found that unmarried, lower education level and poverty are predictors of unsafe abortion [10, 12, 14, 15]. Also, some pregnancy-related factors such as being unaware that abortion is free, unwanted pregnancy, unmet need for modern contraceptive methods, and being unaware that abortion is legal have contributed to inequalities in unsafe abortion [16-19].

In Zambia, part of the government’s solution to reduce unsafe abortion was to make abortion legal by enacting the abortion law 50 years ago [20]. However, despite having one of the most liberal abortion laws in the sub-Saharan African region, more than half of gynaecological admissions and almost one-third of maternal deaths are due to unsafe abortion [21].

Literature suggests that unsafe abortion is pro-poor in most settings [10, 11] but in Zambia there is no evidence of empirical data. As far as literature was searched, no empirical study that has reported socioeconomic inequalities in unsafe abortion in Zambia. Understanding inequalities in unsafe abortion is important to inform policymakers and design possible interventions to avoid unsafe abortion so that no woman is left behind. Thus, we aimed to quantify the socioeconomic inequality in unsafe abortion among women presenting for abortion care services in Lusaka and Copperbelt provinces of Zambia and decompose the determinants contributing to such socioeconomic inequality.

## Methods

### Study design and setting

This cross-sectional study was conducted in nine public hospitals located in Lusaka and Copperbelt provinces of Zambia between August and September 2021. The hospitals were Ndola and Kitwe Teaching Hospitals located in the Copperbelt province, while Women and Newborn, Levy Mwanawasa Teaching, Chipata, Kanyama, Chawama, Matero Chilenje, and Chipatafirst level hospitals are located in Lusaka province. The selection of hospitals from the two provinces was based on the availability of trained health personnel who provide CAC services. These hospitals are largest in the two provinces attend to obstetric and gynecological problems and serves as delivery hospitals for newborns. Also, they serve a variety of clients from different socioeconomic statuses across the two provinces and referrals from other provinces since the two provinces are the most developed in Zambia. The two provinces constitute about 33% of Zambia population.

### Data collection

The data was collected from women who presented for CAC services and were consecutively enrolled in the study. This is because of the sensitive nature of abortions in Zambia and the perceived criminalization. This assisted in attaining the desired sample size within the study period.

We adopted a questionnaire based on literature [7, 22] and Obstetrician/Gynaecologist consultants validated it through a consultative process before piloting it. We piloted the questionnaire in six health facilities in Lusaka on 15 participants, their data was not included in the analysis. The questionnaire was interviewer administered. After piloting, questions were edited and data collectors (nurses and medical students) were trained to understand the questionnaire, study protocol, and collect data. The training was done over five days on the use and configuration of the Open Data Kit (ODK) and ODK collect tool for data collection using android-based mobile tablets and how to submit data to an online Ona server. Data collection was in real-time and uploaded from the health facilities to the server and managed using ODK Aggregate, an intermediate platform for the server. Data was later downloaded into excel format and cleaned.

### Sample size and sampling

We used a single proportion formula to determine the minimum number of women who sort abortion services to be included in our study. We made the following assumptions; prevalence of unsafe abortions was at 25% [23], type 1 error rate of 5%, at 95% confidence intervals. After adjusting for non-response rate (10%) we determined that 318 records would be sufficient. Weighting was done according to the total number of abortion services expected per hospital during the time of the study. Based on the anecdotal data from the hospitals record books of women seeking abortion services, we consecutively recruited the women and the proportion of the calculated sample size was allocated for each hospital as follows: From Women and Newborn Hospital (6.9%), Levy Mwanawasa (14.3%), Kitwe (13.5%) and Ndola (14.3%) Teaching Hospitals. From the first level hospitals: Kanyama (17.2%), Chilenje (6.6%), Chawama (5.3%), Matero (10.1%) and Chipata (11.8%). Eligible participants were identified from the hospital wards dedicated to caring for women with gynaecological emergencies, which is the entry point for women with abortions. Once care was given and the women were stable, the research assistants would approach the women, check for eligibility and discuss study participation with them. Only consenting women were included in the study.

### Measures

#### Outcome variable

In this study, the outcome variable was abortion status which was dichotomous (coded as “0” safe and “1” unsafe). Unsafe abortion was defined based on WHO criteria as the termination of pregnancy carried out by an individual lacking the necessary skills or in an environment below minimal medical standards or both [3].

#### Explanatory variables

Socioeconomic determinants of unsafe abortion, included variables such as demographic characteristics (age, marital, status) socioeconomic (wealth index, employment, residence, education), pregnancy-related characteristics (history of unwanted pregnancy, history of miscarriage, use of modern conceptive methods, awareness of whether abortion is legal in Zambia, awareness of whether hospital offer free abortion care services) as shown in Table 1.

**Table 1:** Social and pregnancy-related determinants of unsafe abortion

| Variable | Operational definition |
| --- | --- |
| Age in years | Age was categorized ($\leq 24$, 25 – 34, $\geq 35$ years) |
| Marital Status | Married, unmarried |
| Education level | Primary or none, secondary, Tertiary |
| Employment status | Employment, unemployed |
| Residence (density) | Low density, medium density, high density |
| Religious denomination | Catholic, Pentecostal, protestants, others |
| Wealth index (quintiles) | Poorest, poorer, middle, richer, richest |
| History of unwanted pregnancy | Yes, no |
| History of miscarriage | Yes, no |
| Use of modern contraceptives | Yes, no |
| Awareness that abortion is legal in Zambia? | Yes, no |
| Awareness of hospitals offered free abortion services | Yes, no |

### Socioeconomic measurement

We used wealth index as a measure of socioeconomic position. Household assets were used to calculate wealth index using principal component analysis (PCA), a standardized method [24]. We assessed socioeconomic status of women using a household asset index as a proxy of socio-economic status [24]. Household assets that were used are a source of drinking water, type of toilet, type of wall for the house, cooking fuel and type of house floor. Also, whether or not a women owned television, radio, mobile phone, land phone, computer, washing machine, fridge, motorcycle, car, house and employed someone, livestock, and agriculture land. Wealth index was constructed using a summation of all weights of included variables generated by PCA after the assumption of Bartlett’s test, Kaiser-Meyer-Olkin measure of sampling adequacy and determinants of matrix correlation were met [25]. Household economic status was constructed using PCA, assuming the input variables were normally distributed since all variables used to construct the PCA were dichotomous. Following PCA, wealth index on a continuous scale was later categorized into five socioeconomic quintiles: the poorest, poorer, middle, richer and richest [26] used for analysis.

### Statistical analysis

Prevalence of unsafe abortion and descriptive characteristics of variables were presented using frequencies and percentages. During the analysis, we adjusted for possible clustering effect due to women were from different hospitals. In this study, socioeconomic inequalities in unsafe abortion were estimated using concentration index (C), and we illustrated it using concentration curve. C was used to estimate the degree of inequality in the distribution of unsafe abortion among women. The plot for the concentration curve demonstrates the cumulative percentage of unsafe abortion on the Y-axis while the X-axis, the cumulative percentage of the women ranked by socioeconomic status starting from the poorest to the richest. Inside the curve is a 45° diagonal line of equality which suggest equal distribution of unsafe abortion. The value of C ranges from -1 to +1 and the value of C=0 implies perfect equality. C’s positive or negative values suggest that inequality in unsafe abortion is disproportionality among the rich or poor women, respectively.

The C has been extensively used as a measure of socioeconomic inequality and decomposition of CI as dependable measure of health inequality [27]. Erreygers-type decomposition analysis of concentration index by determinants of unsafe abortion was conducted, which is a regression-based and quantification of individual explanatory variables contribution to concentration index [27]. A linear additive regression of unsafe abortion (Y) can be expressed as:

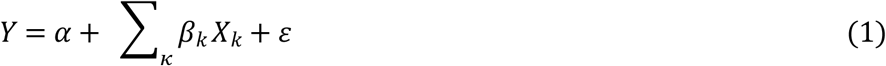

Which can be stated as:

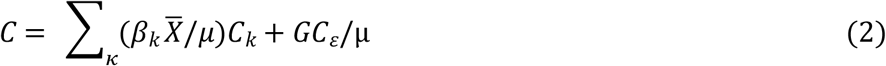

Where Y is the proportion of an unsafe abortion (dichotomous variable) or in case of a continuous variable µ is the mean, k is the determinants, *β*_*k*_ is the coefficient for determinants k from generalized linear regression, for *X*_k_ it is the mean of X for, *C*_k_ is considered as concentration index for *X*_k_, and GCε is given as concentration index for the error term (ε). As presented in equation (2), C is given as weighted sum of the concentration indices for k determinants, and weight for *X*_k_ represents the elasticity of Y with respect to *X*_k_ [27].

In this study, given that unsafe abortion was dichotomous outcome and bounded, as the mean of the variables increase then concentration index becomes smaller and to overcome this challenge corrected concentration index which is specific for binary outcome was calculated. However, Word Bank proposed that from probit model marginal effects can be applied in a non-linear model to restore linear assumption in a decomposition analysis which can be represented as follows:

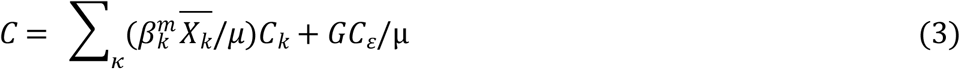

From the probit model, the marginal effect is represented by the coefficient (*β*_*k*_) which give the size of the relationship between the outcome variable and each of the explanatory variables. From equation (2), the component 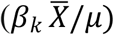 is the elasticity, which is basically the marginal effect when multiplied by the mean of the independent variable and then divided by the mean of the outcome variable. So theoretically, to explain the inequalities in unsafe abortion, this was done by independent variables (contributing factors). *C*_*k*_ in equation (2) represents the concentration indices of the contributing factors that measure inequality and add to the overall concentration index. In this study, we reported relative contribution, which is the how much percentage inequality in the unsafe abortion was attributed to the inequality in the contributing factors which was further adjusted. Lastly, variance inflation factor (VIF) was used to check for multicollinearity among the independent variables with the cut-off point of 1/VIF not more than 0.1. None of the variables in the model suggested multicollinearity. The model included possible interaction terms and none reached any statistical significance [28]. For all analysis the statistical significance level was set at alpha 0.05 (two-tailed). All analyses were conducted with Stata 16.1 (Stata Corp., College Station, Texas, USA).

### Ethical considerations

Ethical approval for this study was sought from University of Zambia Biomedical Research Ethical Committee (reference number: 1852-2021), and further permission was obtained from the Zambia National Health Research Authority, provincial health office, district health office and the senior medical superintendents for each study site. The interviewers introduced themselves and explained the purpose of the study and the hospitals where the study was conducted had no influence on the study. Further, participants were informed that participation was anonymous, purely voluntary, privacy was maintained, information gathered was confidential and at any time they were free to withdraw from the study without affecting their medical care. Written informed consent or assent was obtained from the participants. All the participants were informed that they had chance to leave the study at any time without affecting their medical care.

## Results

### 4.1 Descriptive statistics

In this study, they were 376 women who were eligible participants. After excluding 14 who had missing data on unsafe abortion, 362 were included in the analysis of whom 77 (21.3% [95% CI: 17.6, 26.1]) had unsafe abortion. Generally, for socio-demographic characteristics, majority (22.1%) of the women reported unsafe abortion among those aged 35 years or older and (34.2%) not married. While for socioeconomic characteristics, most (27.3%) had none or primary education, (22.7%) were not employed and (33.3%) in the poorest wealth index quintile. Additionally, unsafe abortion was more prevalent (36.4%) among women who reported a history of unwanted pregnancy, (36.4%) from the Copperbelt province, (34.9%) who reported history of induced abortion, (45.5%) did not know that hospitals offered free abortion care services and (28.4%) did not know that abortion is legal in Zambia Largely. The descriptive results showed that unsafe abortion was more common in the deprived social groups (Table 2).

**Table 2:**
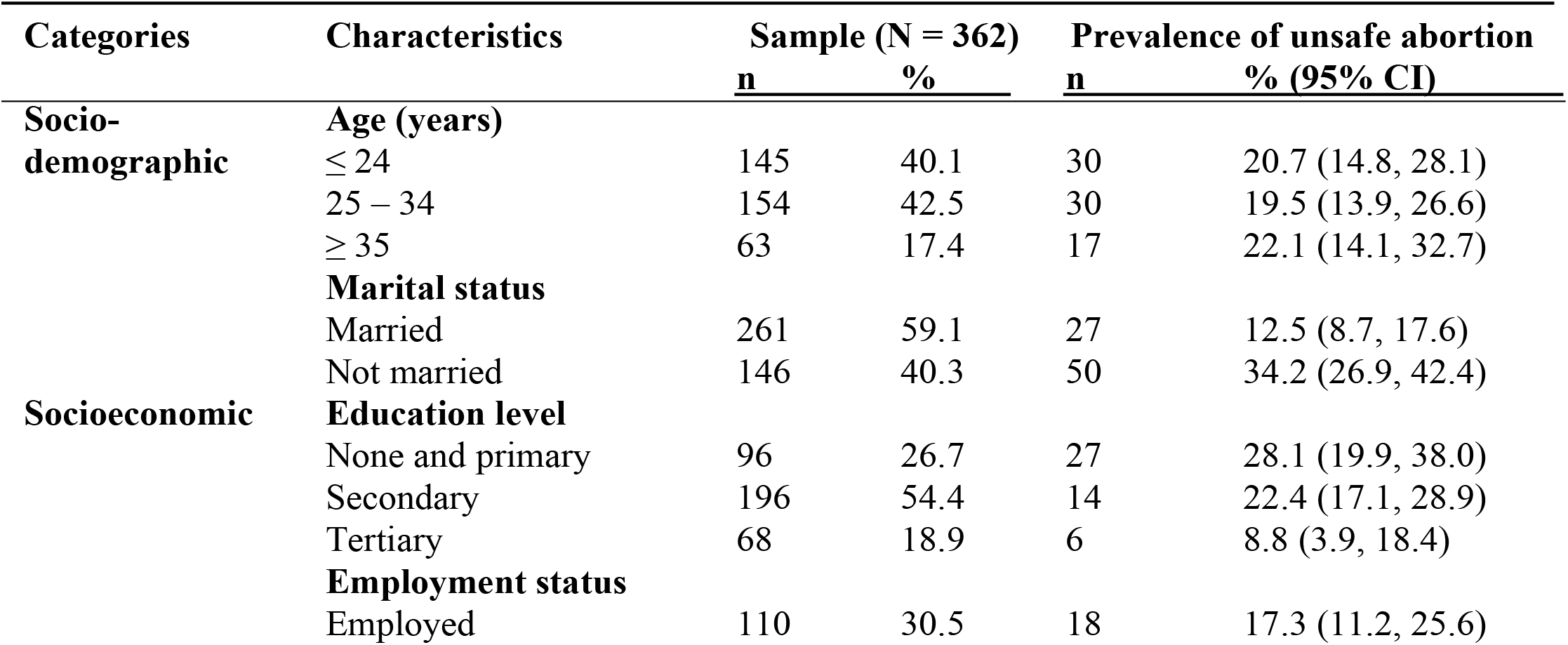
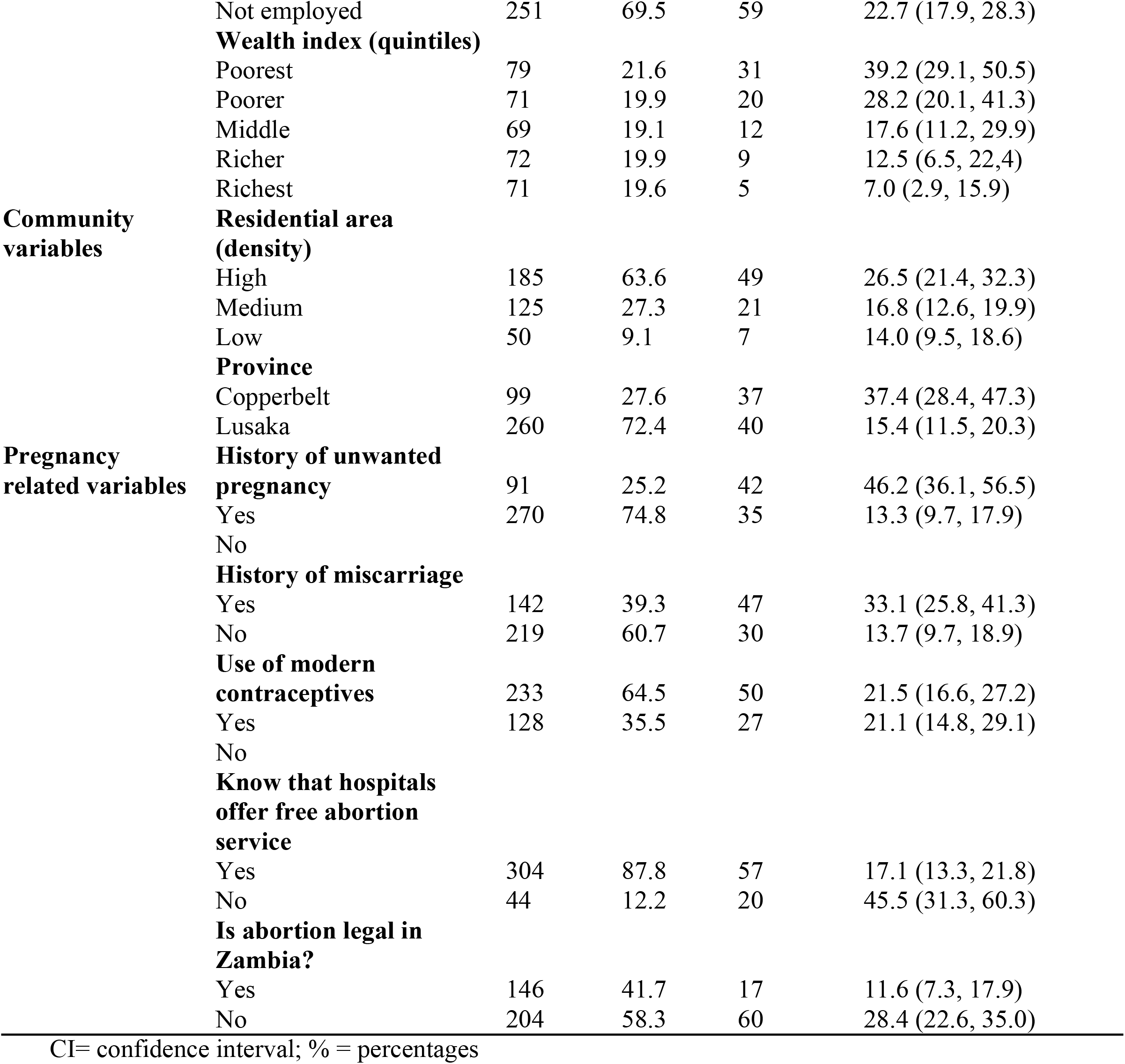
Prevalence of unsafe abortion by women’s characteristics

### 4.2 Decomposition analysis

We observed substantial socioeconomic inequalities in unsafe abortion among women. Table 3 shows a Generalized Linear Regression model based decomposition analysis representing regression coefficient, corrected concentration index, elasticity, contribution and relative percentage contribution of each variable to the total unsafe abortion inequality. In situations where the contribution of a variable is negative, can be considered as *ceteris paribus*, implying that unsafe abortion inequality would be more in the presence of that particular variable but the opposite would occur for a positive contribution.

**Table 3:** Summary of decomposition analysis of socioeconomic inequalities in unsafe abortion in women presenting for abortion care service, 2021 Lusaka and Copperbelt province, Zambia

| Category | Characteristics | Elasticity | $\beta$ | ECI | Contribution to C | Adjusted % Contribution |
| --- | --- | --- | --- | --- | --- | --- |
| Socioeconomic | **Wealth index (ref: poorest)** |  |  |  |  |  |
|  | Poorer | -0.039 | -0.044 | -0.465 | 0.018 | -2.7 |
|  | Middle | -0.133 | -0.089 | 0.008 | -0.001 | 0.3 |
|  | Richer | -0.147 | -0.151 | 0.508 | -0.749 | 12.4 |
|  | Richest | -0.251 | -0.237 | 0.999 | -0.251 | 53.9 |
|  | **Age, years (ref: $\leq 24$)** |  |  |  |  |  |
|  | 25 – 34 | 0.151 | 0.091 | 0.192 | 0.029 | -3.5 |
| | $\geq 35$ | 0.122 | 0.202 | 0.030 | 0.004 | -0.9 |
|  | **Marital status (ref: married)** |  |  |  |  |  |
|  | Single | 0.269 | 1.368 | -0.073 | -0.019 | 6.3 |
|  | **Education level (ref: primary)** |  |  |  |  |  |
|  | Secondary | 0.066 | 0.239 | -0.036 | -0.002 | 2.8 |
|  | Tertiary | -0.023 | -0.240 | 0.714 | -0.0016 | 7.4 |
|  | **Employment status (ref: employed)** |  |  |  |  |  |
|  | Unemployed | 0.029 | -0.183 | -0.232 | -0.007 | 3.7 |
|  | **Residential area (High)** |  |  |  |  |  |
|  | Medium | 0.065 | 0.113 | 0.346 | 0.007 | -1.9 |
|  | Low | 0.222 | 0.326 | 0.144 | 0.072 | -7.7 |
|  | **Province (ref: Copperbelt)** |  |  |  |  |  |
|  | Lusaka | -0.612 | -1.675 | -0.025 | 0.159 | -3.2 |
|  | **Sum** |  |  |  |  | **66.9** |
| **Pregnancy-related variables** | **History of unwanted pregnancy (ref: yes)** |  |  |  |  |  |
|  | No | -0.535 | -1.541 | 0.082 | -0.044 | 5.1 |
|  | **History of miscarriage (ref: yes)** |  |  |  |  |  |
|  | No | -0.196 | -0.351 | 0.014 | -0.003 | 0.7 |
|  | **Use of modern contraceptive (ref: yes)** |  |  |  |  |  |
|  | No | 0.021 | 0.119 | -0.123 | -0.003 | 0.9 |
|  | **Know hospital offered free abortion care services (ref: yes)** |  |  |  |  |  |
|  | No | -0.556 | 0.205 | 0.354 | -0.197 | 11.3 |
|  | **Is abortion legal in Zambia (ref: yes)** |  |  |  |  |  |
|  | No | 0.252 | 0.120 | -0.307 | -0.077 | 8.9 |
|  | **Sum** |  |  |  |  | **26.9** |
| **Total observed** |  |  |  |  |  | **93.8** |
| **Residuals** |  |  |  |  |  | **6.2** |
| **Total** |  |  |  |  |  | **100** |

Notes: C = Concentration Index (Erreygers corrected). \* Residual is the part of socioeconomic related inequality not explained by variables included in the analysis

Women who were from high residential density areas, not employed and had primary education were more concentrated among the poor. Those who knew that abortion is legal in Zambia had positive indices indicating that it was concentrated among rich women. However, for the variable of history of unwanted pregnancy was more an attribute of the poor.

### 4.3 Inequality in unsafe abortion

In this study, C of unsafe abortion was −0.232 (95% CI: − 0.309, − 0.154), suggesting that unsafe abortion was more concentrated among women of low socioeconomic status. The concentration curve of unsafe abortion lays above the 45° line or diagonal line (line of equality) suggesting that unsafe abortion was more among the worse-off women (Figure 1). Table 3 shows the results of decomposition analysis and the C of the independent variables showed that unmarried, secondary education, unemployed, living in Lusaka province, living in high density areas, not using contraceptive methods and unaware that abortion is legal in Zambia were more concentrated among women of lower socioeconomic status. In contrast, age of 25 year and older, tertiary education, living in medium and low density areas, no history of unwanted pregnancy and miscarriage were more common among women of higher socioeconomic status. The categories of variables studied were socioeconomic and pregnant-related variables and these explained 66.9% and 26.9% respectively. About 6.2% was unexplained (residual) inequality. Overall, from the social determinant factors, the major contributors to the observed inequality in unsafe abortion was socioeconomic status (66.6%), marital status (6.3%), education (10.2%) and employment (3.7%). Overall, these variables contributed 66.9% to the total inequality. Furthermore, from pregnancy-related variables history of unwanted pregnancy (5.1%), awareness whether abortion is legal in Zambia (8.9%) and knowledge that hospital offered free abortion services (11.3%). Use of modern contraceptives, history of miscarriage and age 35 years or order were almost equally distributed among women. Other factors such as residence (−9.6%), province (−3.2%) and age (−4.4%) contributed negatively to the inequality. Regarding the contributing variables, it means that given value of contributor Y is y and has a negative (or positive) value, the inequality in unsafe abortion would increase (or decrease) by y% if the contributors was evenly distributed among women of different socioeconomic levels (Table 3).

**Figure 1:**
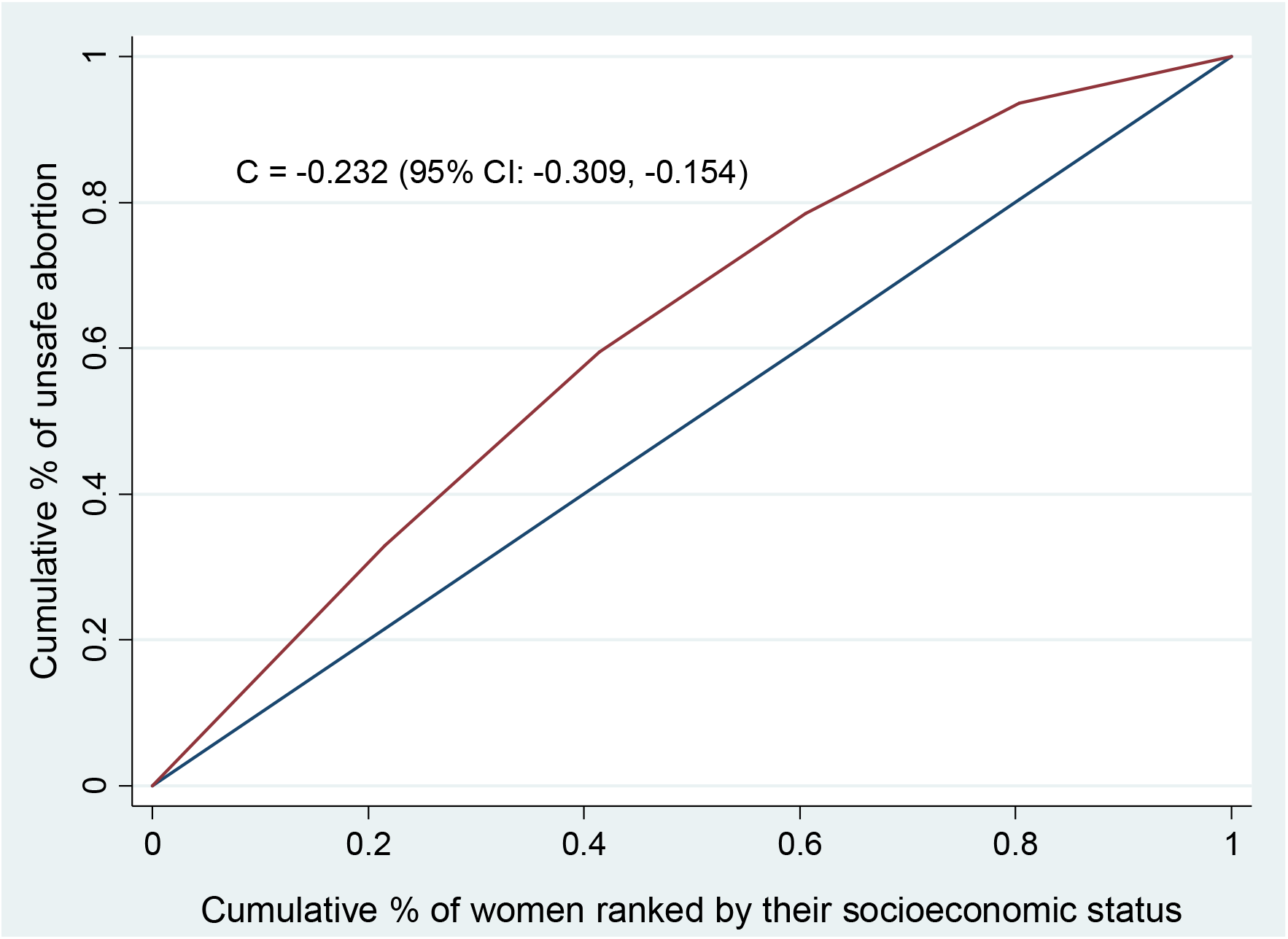
Concentration curve for cumulative proportion of unsafe abortion ranked by wealth index among women who presented for abortion care services in public hospitals in 2020, Lusaka and Copperbelt provinces, Zambia

## Discussion

To the best knowledge of the authors, this is the first study to examine socioeconomic inequalities in unsafe abortion among women presenting for abortion care services in Zambia. The prevalence of unsafe abortion reported in this study was lower than 45% in Ethiopia [22], 36% in Kenya [29] and 67.1% in India [30] but was higher than 16% from Nepal [7]. The differences could be attributed to different abortion policies between countries, access to abortion services, availability of trained CAC providers and different socioeconomic status of the women. The existence of high levels of preventable unsafe abortion in Zambia can exert unprecedented pressure on the already overburdened health service delivery and hinder the achievement of Millennium Development Goal (MDG) number three and increase maternal mortality.

The finding of this study suggest significant socioeconomic inequalities in unsafe abortion since the corrected concentration index was negative (−0.231), implying pro-poor inequality in unsafe abortion among women. Decomposition analysis showed that the major contributors of the unsafe abortion inequality were socioeconomic status, marital status, education, employment, history of unwanted pregnancy, awareness about hospitals that offered free abortion care services and awareness that abortion is legal in Zambia.

In this study about 66.9% of the inequality in unsafe abortion was explained by socioeconomic status. These factors have been shown in many other studies as contributors to the inequalities of unsafe abortion and other health outcomes [11, 18]. This suggests that given socioeconomic status was evenly distributed among women of different socioeconomic groups, unsafe abortion would reduce by 66.9%. Socioeconomic disparities raise differences in access and affordability to many healthcare services culminating in different health outcomes [31, 32]. Although the Zambian economy strive to improve and undergoing major transformation, there are still many people of low socioeconomic status who cannot afford basic health care [33] and this could be a plausible explanation of why unsafe abortion was pro-poor.

We observed that residence has a significant contribution to unsafe abortion inequality. Unsafe abortion was more common among women from high-density residential areas than their counterparts from low residential density areas. High density residential areas are more likely to be poor and previous studies had demonstrated that women who reside in such areas have increased chance of experiencing unsafe abortion [10]. On the other hand, low density areas are associated with wealthy neighborhoods and have better financial capacity and opportunities to health care and other public services [11]. Also, there is a possibility of discrepancy between the number of health facilities and population, so access and adequate services may arise as women from high density residential areas are less likely to access abortion care services. Therefore, policymakers and program implementers should try and reduce the gap between these residential areas.

This study found regional contribution to unsafe abortion inequality. Women from Lusaka province had a negatively contribution to the inequality in unsafe abortion. This finding could be due to differences in access to hospitals, trained CAC providers and different socioeconomic status between the two provinces. In the recent past, there has been less economic development activities in the Copperbelt province due to the sale of copper mines, which was the main economic activity and hub for Zambia economy for many years. This could have affected economic status of many women leaving majority of them in low socioeconomic strata and unable to afford decent basic health care including CAC services.

This study observed that history of unwanted pregnancy contributed to unsafe abortion inequality. Women who had history of unwanted pregnancy were mainly found among those who had unsafe abortion. This finding is in keeping with a previous study that reported that unwanted pregnancy is associated with high likelihood of abortion and unsafe abortion [19]. Unwanted pregnancy may reflect unmet need of modern contraceptives which still is a challenge in most developing countries such as Zambia [18]. This may generate inequality in access and utilization to which women who are poor and of low socioeconomic status are less privileged. In addition, lack of access to abortion care services from public health facilities may predispose women seeking clandestine abortion services from unqualified persons or “wise women” who are more readily available. Hence, acceleration and consolidation of modern contraceptive use may be an urgent need, which has the potential to reduce unwanted pregnancy and disparities among women.

Additionally, awareness whether abortion is legal in Zambia contributed to unsafe abortion among women. Generally, knowledge regarding legality of abortion law enhances utilization of CAC services resulting in reduced unsafe abortion [12, 34]. We found a higher proportion of those who knew that abortion is legal in Zambia in the well-off than among the poor. This suggest that lack of knowledge about legality of abortion in Zambia increases pro-poor unsafe abortion among women. Therefore, policymakers should put in place deliberate measures to publicly enhance information dissemination about the legality of abortion in Zambia especially among the poor women to reduce unsafe abortion. Our finding is in consulates with a studies from Nepal and Ghana which showed that women who were not aware that abortion being legal had increased olds of having unsafe abortion [7, 12].

Regarding knowledge of women whether abortion services are free from public hospital contributed 11.3% of the inequality in unsafe abortion. There was a higher proportion of unsafe abortion among women who were not aware that abortion services are free from public hospitals. Other studies have reported similar findings [11, 35] and a plausible explanation could be that those poor women may fear being turned away resorting to clandestine abortion methods that are unsafe.

In our study, we noted that use of modern contraceptive methods did not contribute much to the inequality in unsafe abortion. Extant literature has suggested that in most resource-poor settings, there is unmet need of modern conceptive among women of reproductive age group associated with unsafe abortion [23, 36]. We suggest qualitative approach could be appropriate to probe into types of contraceptives for both modern and traditional used by women as such issues could be considered sensitive due to stigma and cultural norms. Exploring this issue could help design some appropriate interventions to reduce the unmet need for modern contraceptives. Since all women in this study presented for abortion care services, they never wanted the pregnancy and more than one-third indicated that they were not using contraceptive method or some of those who were using could have been failure of contraceptives. We think a longitudinal study can add more information around this sensitive topic and adding more variables that contribute to the inequality in unsafe abortion.

### Strength and Limitation

Out study had several limitations. First, we did not collect an income variable which some studies have recommended as a socioeconomic ranking variable. However, some surveys have indicated that household assets can be used as a proxy indicator for inequalities and it is sensitive to socioeconomic distribution [25, 37]. Second, we collected data from just two out of 10 provinces and therefore, generation of results may be limited. However, these two provinces are makeup one-third of the Zambia population, with trained CAC provider and information about women seeking abortion care service is readily available.

Third, women were being asked who initiated abortion and from where which was a subjective method of getting information since we could not employ an objective method and this could have led to bias resulting into underestimation of unsafe abortion as women are more likely to under report for fear of apprehension and stigma. Our strengths include this is the first study in Zambia that has explored the socioeconomic inequalities in unsafe abortion and the contributing factors are modifiable and this has highlighted what was previously was not known. Use of the corrected concentration index solved the inherent problem of misestimating the inequality when the outcome variable is bounded.

## Conclusion

Our study has shown a remarkable pro-poor inequality in unsafe abortion exists even within an already resource-poor country. Although the socioeconomic status contributes the greater inequality, even pregnancy-related factors contribute to the inequality in unsafe abortion. Therefore, program implementers and policymakers should take a multi-sectorial approach in mitigating unsafe abortion by empowering women to reduce socioeconomic status gap so that no woman is left behind in accessing free, legal abortion care services regardless of their socioeconomic status.

## Data Availability

The authors will make data available after accepting this manuscript for publication without restrictions.

## Authors’ contributions

Author Contributions: Conceptualization, P.K., M.K.L., and C.C.P.; methodology, P.K., M.K.L.; V.S., validation, P.K., M.K.L., and M.M.; data analysis, P.K; investigation, P.K., M.K.L., C.J., M.M., M.M., C.C.P, B.V., V.S., M.N.M., and M.K.L.; writing-original draft preparation, P.K., M.K.L., and M.M.; writing-review and editing, P.K., M.K.L., M.M., M.M., C.J., C.C.P., M.N.M., and B.V.; research supervision, P.K.; M.K.L., M.M., B.V., V.S., project administration, P.K., M.K.L., M.M., V.S., and B.V. All authors have read and agreed to the published version of the manuscript.

## Institutional Review Board Statement

Ethical approval was obtained from University of Zambia Biomedical Research Ethics Committee (approval number: 1852-2021).

## Informed consent statement

Written informed consent was obtained from all participants involved in the study.

## Data availability statement

The data for this study can be made available upon request from the corresponding author.

## Acknowledgements

The authors sincerely thank International Federation of Gynaecology and Obstetrics for funding this study. The funder had no role in the design of the study, collection, analysis, and interpretation of data or in writing the paper. MM and MKL would like to acknowledge that some of his time is supported by the UNC-UNZA-Wits Partnership for HIV and Women’s Reproductive Health which is funded by the U.S. National Institute’s Health (grant number: D43 TW010558). Also, we acknowledge the effort by the members of the Young Emerging Scientists Zambia who took part in data collection. Further, we acknowledge the support of the Zambia Association of Gynecologist and Obstetrics.

## References

1. Singh S: Global Consequences of Unsafe Abortion. Women’s Health 2010, 6:849–860.

2. Singh S, Maddow-Zimet I: Facility-based treatment for medical complications resulting from unsafe pregnancy termination in the developing world, 2012: a review of evidence from 26 countries. Bjog 2016, 123:1489–1498.

3. WHO: Trends in Maternal Mortality: Estimates by WHO, UNICEF, UNFPA, World Bank Group and the United Nations Population Division, Geneva, Switzerland,. WHO health trends report 2015.

4. Lu C, Cuartas J, Fink G, McCoy D, Liu K, Li Z, Daelmans B, Richter L: Inequalities in early childhood care and development in low/middle-income countries: 2010-2018. BMJ Glob Health 2020, 5:e002314.

5. Diniz SG, d’Oliveira AF, Lansky S: Equity and women’s health services for contraception, abortion and childbirth in Brazil. Reprod Health Matters 2012, 20:94–101.

6. Gil-Lacruz AI, Gil-Lacruz M, Bernal-Cuenca E: Socio-Economic Determinants of Abortion Rates. Sexuality Research and Social Policy 2012, 9:143–152.

7. Yogi A, K.C P, Neupane S: Prevalence and factors associated with abortion and unsafe abortion in Nepal: a nationwide cross-sectional study. BMC Pregnancy and Childbirth 2018, 18.

8. Addor V, Narring F, Michaud PA: Abortion trends 1990-1999 in a Swiss region and determinants of abortion recurrence. Swiss Med Wkly 2003, 133:219–226.

9. Helström L, Zätterström C, Odlind V: Abortion rate and contraceptive practices in immigrant and Swedish adolescents. J Pediatr Adolesc Gynecol 2006, 19:209–213.

10. Fusco CLB: Unsafe Abortion: a serious public health issue in a poverty stricken population. Reprodução & Climatério 2013, 28:2–9.

11. Sousa A, Lozano R, Gakidou E: Exploring the determinants of unsafe abortion: improving the evidence base in Mexico. Health Policy and Planning 2009, 25:300–310.

12. Boah M, Bordotsiah S, Kuurdong S: Predictors of Unsafe Induced Abortion among Women in Ghana. Journal of Pregnancy 2019, 2019.

13. United Nations Department for Economic and Social Information and Policy Analysis: Population Division. World Population Prospects 1995, The 1994 Revision:135.

14. Calvert C, Owolabi OO, Yeung F, Pittrof R, Ganatra B, Tunçalp Ö, Adler AJ, Filippi V: The magnitude and severity of abortion-related morbidity in settings with limited access to abortion services: a systematic review and meta-regression. BMJ Global Health 2018, 3:e000692.

15. Zafar H, Ameer H, Fiaz R, Aleem S, Abid S: Low Socioeconomic Status Leading to Unsafe Abortion-related Complications: A Third-world Country Dilemma. Cureus 2018, 10:e3458–e3458.

16. Appiah-Agyekum NN: Medical abortions among university students in Ghana: implications for reproductive health education and management. Int J Womens Health 2018, 10:515–522.

17. Levandowski BA, Kalilani-Phiri L, Kachale F, Awah P, Kangaude G, Mhango C: Investigating social consequences of unwanted pregnancy and unsafe abortion in Malawi: the role of stigma. Int J Gynaecol Obstet 2012, 118 Suppl 2:S167–171.

18. Rasch V, Silberschmidt M, McHumvu Y, Mmary V: Adolescent girls with illegally induced abortion Dar es Salaam: The discrepancy between sexual behaviour and lack of access to contraception. Reproductive Health Matters 2000, 8:52–62.

19. Tesfaye G, Hambisa MT, Semahegn A: Induced Abortion and Associated Factors in Health Facilities of Guraghe Zone, Southern Ethiopia. Journal of Pregnancy 2014, 2014:295732.

20. Zambia GotRo: Termination of Pregnancy Act. Ministry of Legal Affairs;. 1972.

21. Macha S, Muyuni M, Nkonde S, Faúndes A: Increasing access to legal termination of pregnancy and postabortion contraception at the University Teaching Hospital, Lusaka, Zambia. International Journal of Gynecology & Obstetrics 2014, 126:S49–S51.

22. Temesgen MA, Maru NM, Reddy PS: Proportion of Unsafe Abortion and Associated Factors among Women Who Need Abortion Services in Health Facilities of Dessie Town, Ethiopia. In.; 2017

23. Haddad LB, Nour NM: Unsafe abortion: unnecessary maternal mortality. Reviews in obstetrics & gynecology 2009, 2:122–126.

24. Rutstein S: Steps to constructing the new DHS wealth index. Rockville, MD 2015, ICF International.

25. Vyas S, Kumaranayake L: Constructing socio-economic status indices: how to use principal components analysis. Health Policy Plan 2006, 21:459–468.

26. Howe LD, Hargreaves JR, Huttly SRA: Issues in the construction of wealth indices for the measurement of socio-economic position in low-income countries. Emerging Themes in Epidemiology 2008, 5:3.

27. Wagstaff A: The bounds of the concentration index when the variable of interest is binary, with an application to immunization inequality. Health Econ 2005, 14:429–432.

28. Sommet N, & Morselli, D. (2021): Keep Calm and Learn Multilevel Linear Modeling: A Three-Step Procedure Using SPSS, Stata, R, and Mplus. International Review of Social Psychology 2021, 34:24.

29. Oyieke JB, Obore S, Kigondu CS: Millennium development goal 5: a review of maternal mortality at the Kenyatta National Hospital, Nairobi. East Afr Med J 2006, 83:4–9.

30. Yokoe R, Rowe R, Choudhury SS, Rani A, Zahir F, Nair M: Unsafe abortion and abortion-related death among 1.8 million women in India. BMJ global health 2019, 4:e001491–e001491.

31. Becker G, Newsom E: Socioeconomic status and dissatisfaction with health care among chronically ill African Americans. American journal of public health 2003, 93:742–748.

32. McMaughan DJ, Oloruntoba O, Smith ML: Socioeconomic Status and Access to Healthcare: Interrelated Drivers for Healthy Aging. Frontiers in public health 2020, 8:231–231.

33. United Nations: Inequality in a Rapid Changing World: World Social Report. Department for Economic and Social Affairs 2020,372.

34. Latt SM, Milner A, Kavanagh A: Abortion laws reform may reduce maternal mortality: an ecological study in 162 countries. BMC Women’s Health 2019, 19:1.

35. Ganatra B, Hirve S: Induced abortions among adolescent women in rural Maharashtra, India. Reprod Health Matters 2002, 10:76–85.

36. Munakampe MN, Zulu JM, Michelo C: Contraception and abortion knowledge, attitudes and practices among adolescents from low and middle-income countries: a systematic review. BMC Health Services Research 2018, 18:909.

37. Gatimu SM, John TW: Socioeconomic inequalities in hypertension in Kenya: a decomposition analysis of 2015 Kenya STEPwise survey on non-communicable diseases risk factors. International Journal for Equity in Health 2020, 19:213.

